# Application and Significance of SIRVB Model in Analyzing COVID-19 Dynamics

**DOI:** 10.1101/2024.09.21.24314129

**Authors:** Pavithra Ariyaratne, Lumbini P. Ramasinghe, Johathan S. Ayyash, Tyler M. Kelley, Terry A. Plant-Collins, Logan W. Shinkle, Aoife M. Zuercher, Jixin Chen

**Affiliations:** Department of Chemistry and Biochemistry, Nanoscale & Quantum Phenomena Institute, Ohio University, Athens Ohio 45701

**Keywords:** SIR, SIRV, SIRVB, COVID kinetic models, global data analysis, undergraduate teaching

## Abstract

In the summer of 2024, COVID-19 positive cases spiked in many countries, but it is no longer a deadly pandemic thanks to global herd immunity to the SARS-CoV-2 viruses. In our physical chemistry lab in spring 2024, students practice kinetic models, SIR (Susceptible, Infected, and Recovered) and SIRV (Susceptible, Infected, Recovered, Vaccinated) using COVID-19 positive cases and vaccination data from World Health Organization (WHO). In this report, we further introduce virus breakthrough to the existing model updating it the SIRVB (Susceptible, Infectious, Recovered, Vaccinated, Breakthrough) model. We believe this is the simplest model possible to explain the COVID-19 kinetics in all countries in the past four years. Parameters obtained from such practice correlate with many indices of different countries. These models and parameters have significant value to researchers and policymakers in predicting the stages of future outbreaks of infectious diseases.

**TOC:** 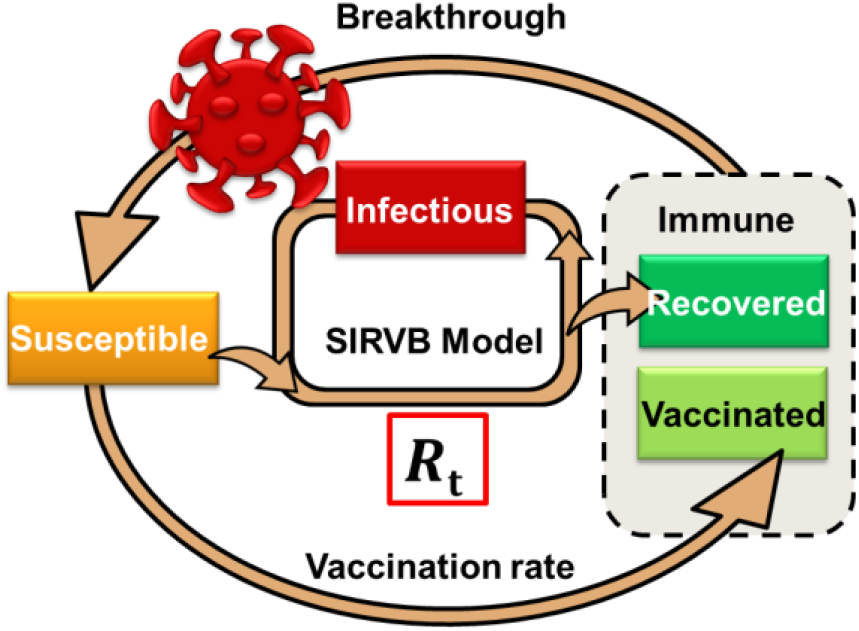

## Introduction

“Pandemic” would have been just a word in modern times, if we all had not experienced the true sense of it until COVID-19 happened. The novel coronavirus, caused by virus SARS-CoV-2, first appeared in Wuhan, China in December 2019 ^1^. The World Health Organization (WHO) announced it a pandemic named COVID-19 in March 2020 and released the alarm in May 2023 ^2,3^. During these three years and until this report in the summer of 2024, over 700 million people around the world have been infected with this virus, and over 7 million people died, with an unknown but estimated much larger number of excess deaths by WHO ^4–6^. Among those, over 1 million deaths in the USA ^7^, indicating the impact of COVID-19 on the country, which spends the highest percentage on healthcare from GDP ^8^. Therefore, it is an interesting study for students to explore how fast COVID-19 spread and recovery using kinetic models.

The COVID-19 pandemic data has been used in our physical chemistry teaching lab course since 2021. In early 2020, when the world was exposed to COVID-19, the kinetic models were used by epidemiologists to predict the impact of COVID-19, way before the world had realized it ^9,10^. The basic reproduction number, *R*_0_, meaning how many people can get infected by one infected person of COVID-19, was estimated ∼3.0 in the early stage ^11,12^, higher than seasonal influenza, which is around 0.9-2.1 ^13^. The incubation period of the SARS-CoV-2 virus was also higher than that of the common flu and society had no natural immunity as it was a novel virus type ^14^. Until vaccines were invented and properly distributed, the epidemiologists suggested that the best way to slow down the spreading was by enforcing social distancing ^15,16^. Throughout the COVID–19 peak period, kinetic models were used as a main tool for decision making ^16^. After 4 years since its first outbreak, positive cases are still reported worldwide daily ^17^, with different variants emerging. Luckily, it is no longer considered deadly due to the cumulative herd immunity achieved by the world. Herd immunity is the point at which the amount of susceptible people is less than 1/*R*_0_ of the total population ^18^. Vaccination and getting infected are the two ways to achieve herd immunity. We are interested in analyzing which country takes which way.

Our in-class practice of COVID-19 kinetics analysis began in 2021, it was introduced to teach students how to do kinetic model analysis using an Excel spreadsheet and enabled them to perform lab work remotely ^12^. Since then, SIR (Susceptible, Infectious, and Recovered) and SIRV (Susceptible, Infectious, Recovered, Vaccinated) models have been used. The SIR model was originally developed by biochemist William Ogilvy Kermack and physician and epidemiologist Anderson Grey McKendrick in 1927 as a meaning to use the simplest possible mathematical model to predict spreading speed ^19^. Modifications have been added over the past years ^20^, such as the SIRV model ^21^. In the past three years, we have been performing this lab experiment, with each year exploring more data. Our first lab in 2021 spring focused on two states’ data in the USA ^12^, and our second lab in 2022 focused on eight states in the USA ^22^. Then our 3^rd^ lab in 2023 focused on different countries across all continents ^23^. This year, in the spring 2024 lab, the COVID-19 data was downloaded from the Our World in Data website ^24,25^. Since the SIR and SIRV models showed difficulty in explaining the reproduction number in the later stages, we are introducing the SIRVB (Susceptible, Infectious, Recovered, Vaccinated, Breakthrough) model to analyze the COVID-19 data. Finding the next simplest model is an essential intention in research and education. It also has practical significance for policymakers to predict stages and to suggest proper social restrictions. The key achievement of these models is to obtain a central parameter *R*_t_, real-time reproduction number, that is independent of testing and can be predicted from social parameters that are readily available ^26^. We believe that the stringency index ^24^ and *R*_0_ alone are enough to estimate *R*_t_ values in these models, providing a fast estimation of COVID-19 spreading speed.

## Method

Using Microsoft Excel and SIR or SIRV model (**Figures 1a, 1b**) with the forward Euler method to extract the real-time effective reproduction number *R*_E_ and real-time reproduction number *R*_t_ has been explained in our previous publications ^12,22,23^. The SIRVB model used in this report has the same structure as the SIRV model with one additional infection pathway (**Figure 1c**). The once immune people may still get infected with a breakthrough rate simplified to a single number *b*. This number is difficult to find and may change over time after the day when a person has recovered or been vaccinated ^27^. The meanings of the parameters are shown in **Figure 1**, where the boxes represent the number of people in each category, and the variables on the arrows represent the rate of transfer to another category. The discrete SIR and SIRV models have been detailed and explained in our previous publications ^12,22,23^. We are introducing the data analysis procedure of the Discrete SIRVB model (**Figure 1c**) as follows. The analysis can be done in Microsoft Excel, which is relatively time-consuming and is suitable if a relatively larger class shares the load. In this report, we have coded MATLAB codes to speed up the process by automatizing the Excel process we have been doing before.

**Figure 1.**
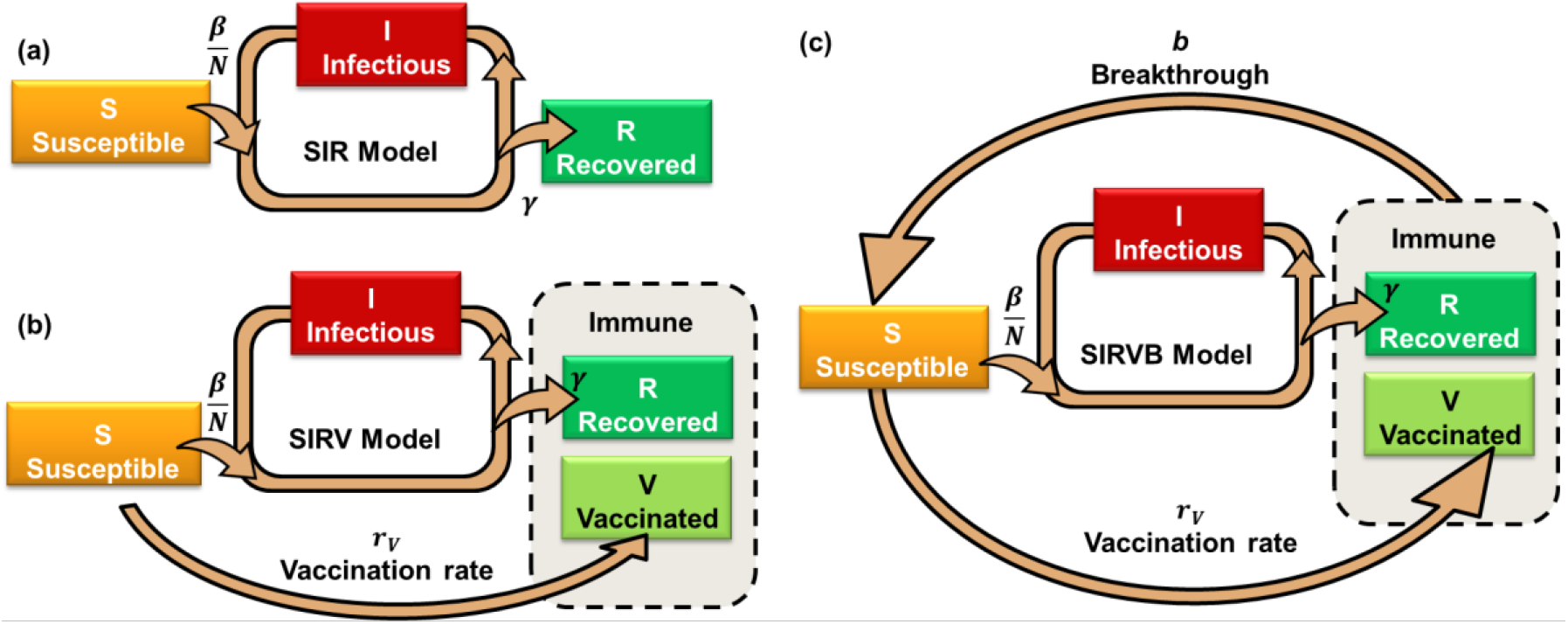
Scheme of (a) SIR, (b) SIRV, and (c) SIRVB models.

Briefly, from the raw data downloaded from Our World in Data ^24^, we selected the smoothed new tested positive cases (*n*) per million population data with an additional 20-day window Gaussian smooth, and fully vaccinated (V) per million population data to analyze the reproduction numbers over time. We set the discrete time step Δ*t* = 1 day, whose effect on convergence is simulated to be properly small for a million people ^12,22,23^. Thus, the initial values are set to be total population *N* = 1,000,000, *S* = *N*, daily new case *n* = 0, active infectious population *I* = 1, recovered population *R* = 0, vaccinated population *V* = 0, infection rate constant *β* = 0 (day^-1^), recovery rate constant *γ* = 0.2 (day^-1^), vaccination rate constant *r*_v_ = 0 (day^-1^), and breakthrough rate constant *b* = 0.2. *N, γ*, and *b* are simplified to be fixed values throughout the calculations, *n* and *V* are from the raw data, and the rest are calculated variables ^12,22,23^. The average breakthrough rate value of the *R*+*V* population is difficult to find, although it is definitely smaller than 1/*R*_0_, otherwise, the pandemic would have never ended. As an example, we simplify the value to be fixed over time, *b* = 20% based on the literature values,^27–29^ and a few different values will be tested later.

These models resemble the logistics problem of self-pumped water/current flows and can be solved mathematically using various methods, among which we will use the Euler method. According to the Euler forward method, at time *t* = *i*, the *i*^th^ day after day 1 of choice from the raw data, on Jan 5^th^, 2020, the second-order “rate constant” of infection is calculated from the number of new cases *n*_*i*_ from the raw data.

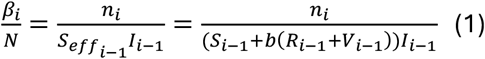

The daily fully vaccinated people *v*_*i*_ can be directly pulled out from the raw data or calculated from the accumulated cases of the fully vaccinated population *V*,

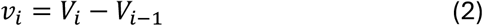

Thus, each day, the susceptible population is reduced by the newly infected and vaccinated population. They are simplified to be no overlap in this report instead of an overlap assumed in the previous classes

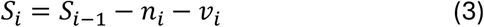

The infectious population gaining from daily infection and losing to daily recovery,

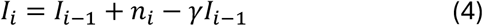

And the recovered population,

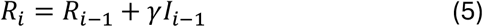

During the calculation, *S* and *I* are kept >=1, and *R* is kept <=*N*-*S-I-V* to avoid zeros in the denominators.

Finally, the real-time effective reproduction number and the actual reproduction number are calculated,

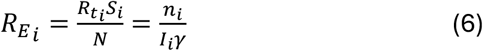

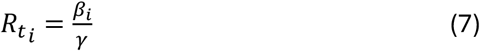

These two curves are then smoothed using the moving median method (15 days) to exclude the spikes caused by irregular data collection.

The purpose of disease control is to reduce the effective reproduction number closer to 1 to flattening-the-curve (FTC) ^30^, or keep it <1 long enough for the epidemic to fade. For a set of experimental data, *R*_E_ values are the same, and *R*_t_ values are dependent on the three models in **Figure 1**. If no action is taken and no virus mutation, *R*_t_ is expected to remain constant throughout the pandemic. With social regulations, *R*_t_ is expected to be inversely proportional to social distance and thus, they carry very useful information ^26^. It can be used to quantify the overall effect of government stringency and social restrictions. Thus, the calculated values in the SIRVB model are further analyzed and correlated to various social data and indices, to find the significance of the SIRVB model in predicting and evaluating COVID-19 kinetics.

## Results and discussion

Assuming a basic reproduction number *R*_0_ = 3.0 for COVID-19, the SIR model predicts that herd immunity will be achieved in 45 days with 1-1/*R*_0_ = 67% of the population infected and recovered if no restrictions are carried out in each million population (**Figure 2**). The infection period can be roughly described as early stage/phase (left part in **Figure 2**), middle stage (the sand-clock region), and later stage (right part). *R*_t_ remains constant in all three stages, similar to the rate constants in a chemical reaction, and *R*_E_ decreases over time. Herd immunity is reached at *R*_E_ = 1. This 2-month simulated lifetime is way faster than the > 3-year pandemic. This is because social regulations have reduced the otherwise constant *R*_t_ thus the spreading is slowed down, a social practice named flattening-the-curve (FTC) ^30^.

**Figure 2.**
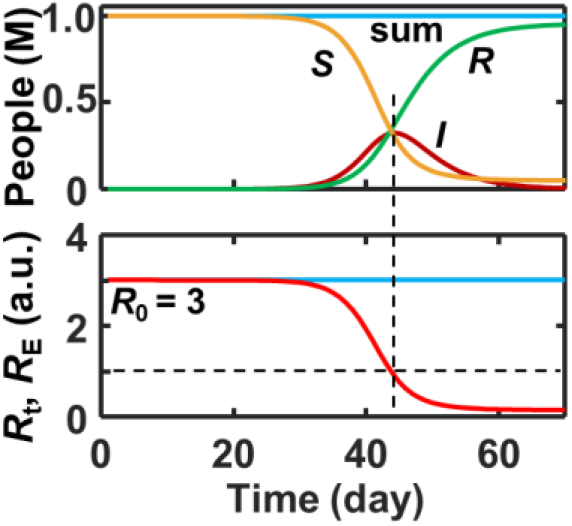
Simulation of SIR model with *R*_t_ = *R*_0_ = 3.0. Detail algorithm available in the literature and an Excel file running the simulation can be found in our previous publications.^12,22,23^

Our models consistently pulled out the effective reproduction number *R*_E_ from the raw data Mathieu *et al*. published on Our World in Data, almost exactly the same but a little noisier than what they had calculated (**Figure 3**).^24^ However, the SIR model ignores the vaccination, and the SIRV ignores the breakthrough, thus both give unreasonable estimations of *R*_t_ at later stages of the pandemic, which are supposed to return to *R*_0_ when social restrictions have been removed, as indicated by the social stringency index returning to zero after the pandemic ^24^.

**Figure 3.**
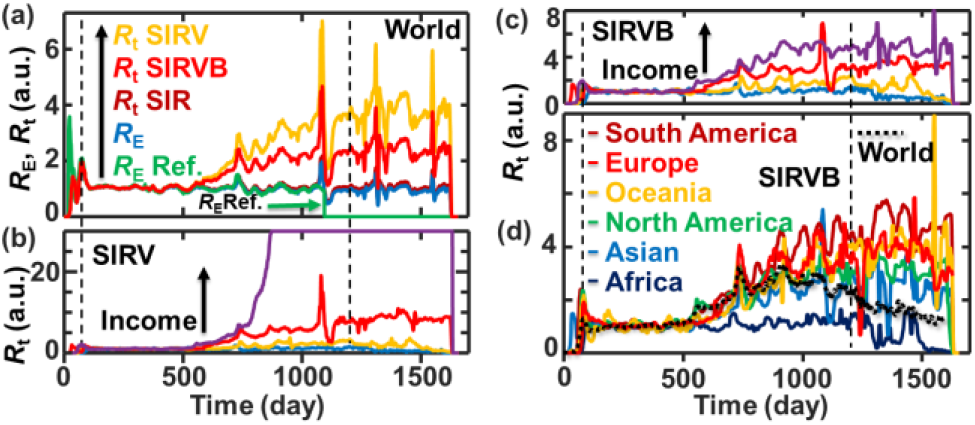
Reproduction number analysis of the world average data grouped and summarized in the raw data by Mathieu *et al*.^24^ (a) *R*_E_ and *R*_t_ values of different models for the world average data. The reference *R*_E_ values is taken from the raw data. *R*_t_ values of (b) SIRV model, (c) SIRVB model for the average of four groups of countries ordered from low to high incomes, and (d) SIRVB model for the countries in the six continents. Data are smoothed. The dashed lines are when WHO announced the start (March 11, 2020) and the end of the pandemic (May 5, 2023).

In these three models, the *n* and *V* values are fixed from the testing data and vaccination data reported to WHO. Fixed *n* values and the *γ* value yield the same *I* values for the three models and thus the same effective reproduction numbers *R*_E_ (**Equation 6**). These values are consistent with what Mathieu *et al*. have calculated (**Figure 3a**). The true *I* values are an epidemiological and statistical challenge to find. The consistency between our *R*_E_ values and those reported by Mathieu *et al*. suggested that we have chosen a similar *γ* value with a proper smooth method and analysis procedure/algorithm.

The three models, SIR, SIRV, and SIRVB, calculate the *S* and *R* populations very differently, yielding very different *R*_t_ values (**Figure 3a**). The SIR and SIRV models have difficulties explaining the *R*_t_ values of the later stages, namely 600-1600 days after the first day of the raw data (Jan 5^th^, 2020). During this period, social restrictions have been lifted and stringency has lowered in most countries. Thus, the *R*_t_ values are supposed to be similar to *R*_0_ if the mutations have a similar *R*_0_ or raised to a plateau if they have a larger *R*_0_.

The SIR model ignores the effect of vaccination in reducing susceptible populations, and typically limited testing may have lower estimated the *I* population, thus, it yields a *R*_t_ very close to *R*_E_ over the whole pandemic and after. We have little interest in using the SIR model to explain the raw data.

The SIRV model seems to work properly at first glance for the world average data with reasonable shape and a reasonable *R*_t_ plateau value ∼4, but it produces an unreasonably large *R*_t_ in the later stages for the counties, especially the developed countries that have a high vaccination rate (**Figure 3b**). This is reasonable because, in the SIRV model, *S* can reach 0 and even negative if not restricted, especially when both vaccination and testing rates are high. *R*_t_ and *β* will go to infinity if S = 0. Our analysis in **Figure 3b** has already restricted *S* and *I* to be >1 and maintained the sum of *S, I, R, V* = *N* but still has trouble. Thus, the SIRV model is not suitable for these countries in the late phase of the COVID-19 pandemic. In our past practice, we have assigned an overlap of the *R* and *V* population but the over-shooting of *R*_t_ still existed in the later stage.

The SIRVB model correctly reflects these expectations in both the world average and most individual countries. However, a dependence on incomes (**Figure 3c**) and continents (**Figure 3d**) is observed. Individual countries also show different *R*_t_ plateau values. Africa data do not have an uprising plateau in the later phase, which is particularly different from the other continents. We believe this is because of the low testing ratio in developing countries ^31,32^. Low statistics will directly affect the calculation of the *R*_t_ and *R*_E_ values. It has been reported that getting COVID data from some countries in Africa has been challenging ^33^, thus we suspect the difference between Africa and other continents is a low statistical problem of the raw data. The average *R*_t_ curve of all 242 countries listed in the raw data is consistently aligned with the average of the curves in **Figure 3c** and **Figure 3d**. The decay of *R*_t_ on the average curves after 900 days is because many countries stop reporting test results, especially after the pandemic. We assume that vaccination data are more accurate because it is easier to track than the other factors thus, we believe uncertainty in vaccination is not the major source of error in this model. The breakthrough *b* value potentially has a big effect on the model and thus is further analyzed.

By changing the average breakthrough rate *b* in the SIRVB model, the equilibrium *R*_t_ plateau can be adjusted (**Figures 4a-4d, Figure 3c**). The average breakthrough rate of COVID-19 in immune people (*V*+*R*) must be <1/*R*_0_ for the pandemic to end. But *b*>0.3 (30%) can be used in the SIRVB model. The plateau *R*_t_ values decrease over the increase of the *b* value, and at *b* = 1, *R*_t_ converges to *R*_E_. Note, the SIRV model could have provided a reasonable plateau of *R*_t_ if we had factored in ratios of *S, I, R*, and *V* to their hidden true values and/or a sophistic overlap model of different populations.

**Figure 4.**
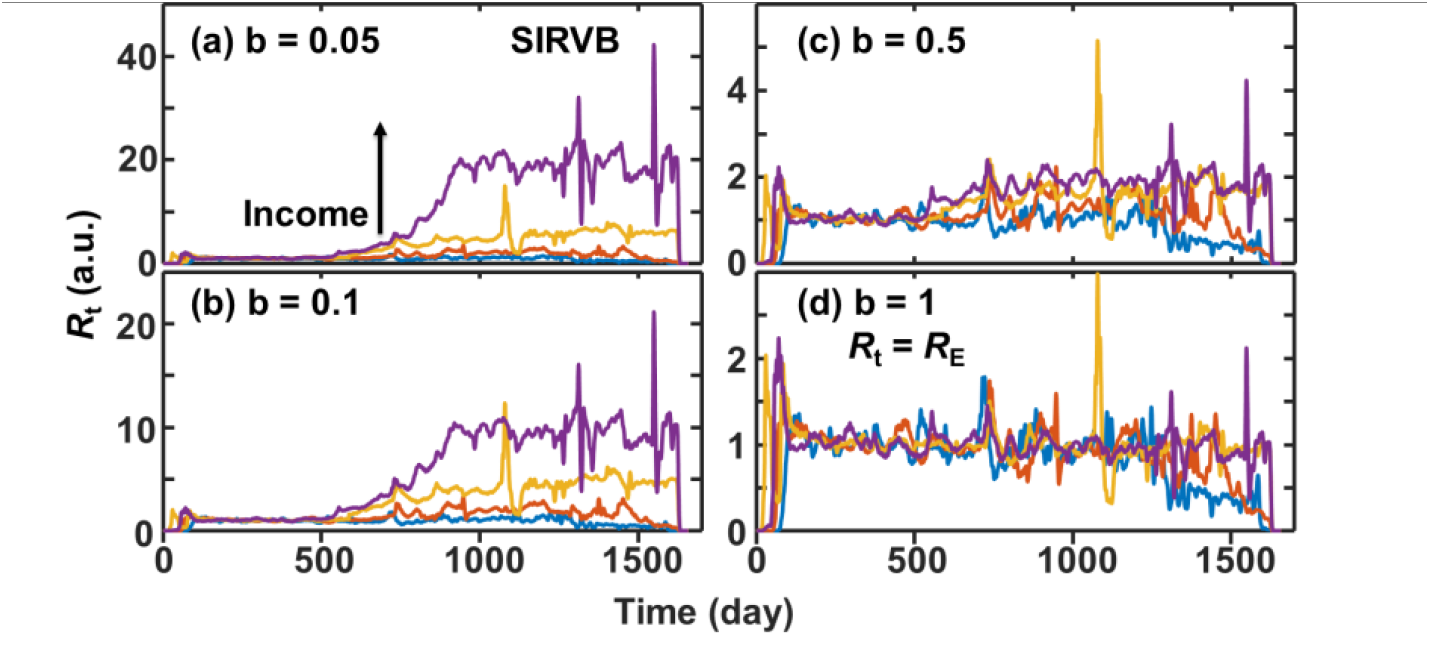
(a-d) The effect of breakthrough rate on the calculations of *R*_t_ values in the SIRVB model. The four groups of countries in the raw data with low to high incomes are used as examples. Curves of *b* = 0.2 have been shown in **Figure 3c**.

The SIRVB model is more straightforward than the SIRV model in that it simplifies all complicated factors into a single parameter *b* and maintains the other values unchanged. Because the protection/immune effect of both infected and vaccinated fades over time ^34–36^, we select *b* = 0.2, a 20% breakthrough rate in the next analysis based on estimations in the literature ^27–29^. In the SIRV model, the cumulative (*I*γ*)_cumu_ = *R*. Introducing a breakthrough rate in the SIRVB model allows (*I*\**γ*) _cumu_ > *R* or even the total population *N* while maintaining *S*+*I*+*R*+*V* =*N* over time. This *b* = 0.2 setting generates *R*_t_ values in 2-4 after 1000 days on the average data of the world and five continents (**Figure 3**). These values are consistent with the estimations of *R*_0_∼3 in the literature ^11^. with no need to assume much bigger *R*_0_ values for the later mutations of SARS-CoV-2. It has been challenging to find the *R*_0_ values for different virus strings ^11,37^. Thus, keeping *R*_0_ the same in a model can be strategically beneficial until solid data are obtained to update its value.

To test the significance of the SIRVB model, we extracted several parameters for each country from the raw data as well as other reports on the Our World in Data website and compared them to the results obtained from the SIRVB model ^38^.

First, *R*_t_ can be used to confirm/detect herd immunity status with high-quality global or local data, e.g. one city. This is because unlike *R*_E,_ which changes over time even if no action is taken, *R*_t_ is a constant only dependent on the nature of the virus and the intensity of social interactions ^39^. Thus, when social stringency is back to normal, *R*_t_ should be close to *R*_0_, which can be used to back-calculate the true immune populations and judge or predict the herd immunity status. For after-pandemic analysis of the raw data, we can simply use the testing data and find the turning point of *R*_t_ at the beginning of the later stage *R*_t_ plateau, which is ∼900 days for the data shown in **Figures 3** and **4**. We propose to use this date as the day of herd immunity i.e. ∼67% of the population is truly immune to SARS-CoV-2 at the middle point of the sand clock region in **Figure 2**.

The consistency of this assignment with other parameters is found in many countries, e.g., for France data (**Figure 5a**). *R*_t_ is calculated from the raw data (**Figure 5a**), and the herd immunity time of France is assigned using *R*_t_ at ∼800 days (**Figure 5b**). This is significantly earlier than the 900 days of the world data. This earliness makes sense because of the fast and high vaccination rate of France compared to the rest of the world, reaching ∼80% at this time (**Figure 5a**). It is also consistent with the time of the biggest wave in France (**Figure 5a**), right before the government relieved social restrictions, and ∼65% of total excess mortality due to COVID-19, which is estimated in the raw data by Mathieu *et al*.^24^

**Figure 5.**
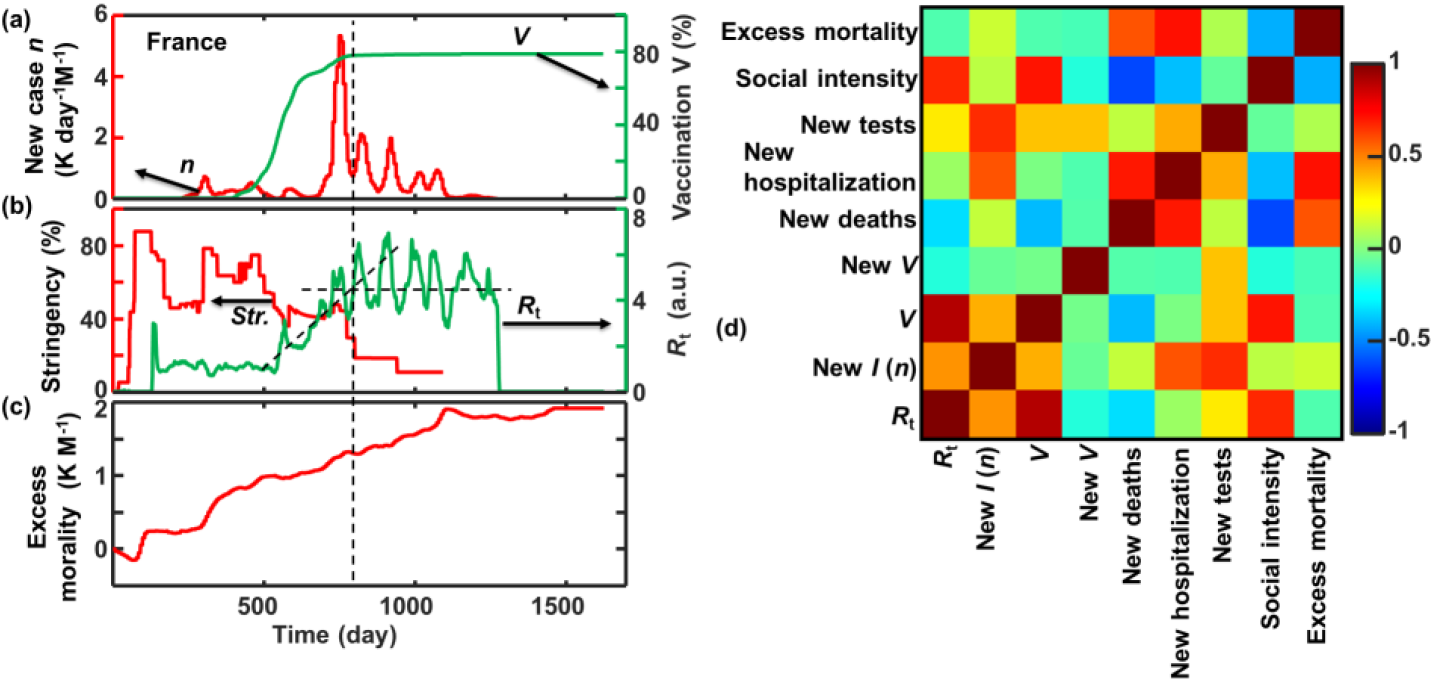
Example data analysis of France. (a) Raw data of *n* and *V*. (b) Social stringency estimated by Mathieu *et al*.^24^ in the raw data and *R*_t_ calculated in this report. Dashed lines indicate our estimation of the herd immune date. (c) Excess mortality estimated in the raw data. (d) Correlation of *R*_t_ with other time dependent parameters for COVID-19 in days 1-1200. If a curve goes up with the increase of another curve, the correlation will be positive, 0 if no correlation, and negative if goes down with the increase of another curve.

Zoom in France’s data and correlate *R*_t_ with other time-resolved raw data, we can also find reasonable correlations and anti-correlations of the evolution of *R*_t_ with other time-resolved data (**Figure 5d**). *R*_t_ is calculated from *n* and *V*, and a positive correlation is observed for the France data. *R*_t_ has a stronger correlation with *V* because social recovery is timely correlated with vaccination in France (**Figures 5a, 5b**). *R*_t_ has low to no correlation with new vaccinations, new deaths, new hospitalizations, and new tests. These low correlations make sense because these parameters are indirectly affect social behavior and may have a delay effect, thus the simple correlation calculation cannot correctly reflect their true correlations.

The positive correlations between *R*_t_, *n* (new daily *I*), *V*, new tests, and the social intensity obtained (inversed) from stringency in the raw data estimated by Mathieu *et al*.,^24^ beautifully explain the flattening-the-curve practice. The same data can be seen from the anti-correlation between *R*_t_ and stringency in **Figure 5b**. If a community halves social intensity/frequency/”temperature” by reducing activities or increasing social distance, it halves the disease transmission and thus halves the *R*_t_. The weak correlations of *R*_t_ with other parameters, especially new *V* and new deaths are good signs because *R*_t_ is a constant in the SIRVB model if there are no social intensity changes. Because of the large *R*_0_ value of COVID-19, it takes less than two months for it to infect the whole population at normal social intensity. Thus, assigning the turning point of the *R*_t_ curve as the herd-immunity date is a reasonable choice.

The other significant correlations or anti-correlations in France data (**Figure 5d**) can be explained. The positive correlations between *n*, new hospitalization, and new tests are expected. The correlations of *V* with many parameters are due to the time correlation not because of causality correlation as indicated by the weak correlation of the new *V* (differential) column. New deaths, new hospitalizations, and new tests are correlated with each other, which are expected. They are also expected to be anti-correlated with social intensity when people adjust their social activities with the correct information of the spreading status. New tests have no effect on true *R*_t_ in the model, but it changes the value of *R*_t_ and social behaviors, so its positive correlations with *R*_t_ and other parameters are observed. But its weak to no anticorrosion with social intensity is surprising. Even if we expect a stronger anticorrelation, this weakening could be due to the wave structure of the new cases and a delay in social responses. Finally, in this figure, the excess mortality is positively correlated with *n*, new death, new hospitalization, and new tests, and anticorrelated with social intensity, which is very reasonable and expected. From the above analysis, we can conclude that the France data is statistically significant and well-behaved in the framework of the SIRVB model.

The *R*_t_ curves of the world average and France both have the laydown ‘5’ shape so do many other countries. Thus, we use this shape to fit data from all countries using a fitting algorithm named “jcfit”, which we have previously developed (**Figure 6a**).^40^ In countries with high testing and vaccination rates, the summation of the infected and vaccinated population (assuming no overlap for simplicity of the model) has exceeded the total population, for example, France. Thus, the *R*_t_ in the later phase is controlled by the breakthrough factor at *R*_t_ ≈ 1/*b*. The turning point occurs on the 802 days. The world average contains data from countries that have low testing and vaccination rates, thus all *R*_t_ < 1/*b*. The global turning point (*t*_4_) is observed on the 940 day (Aug. 2, 2022), 276 days before WHO announced the end of the pandemic^41^ (May 5, 2023), about the average time between the last three big global waves. The slope between *t*_3_ and *t*_4_ reflects the social recovery after the most significant peak of COVID-19 in the area and is correlated with the release of stringency. We assign Aug. 2, 2022, the global herd immunity day on the SIRVB model.

**Figure 6.**
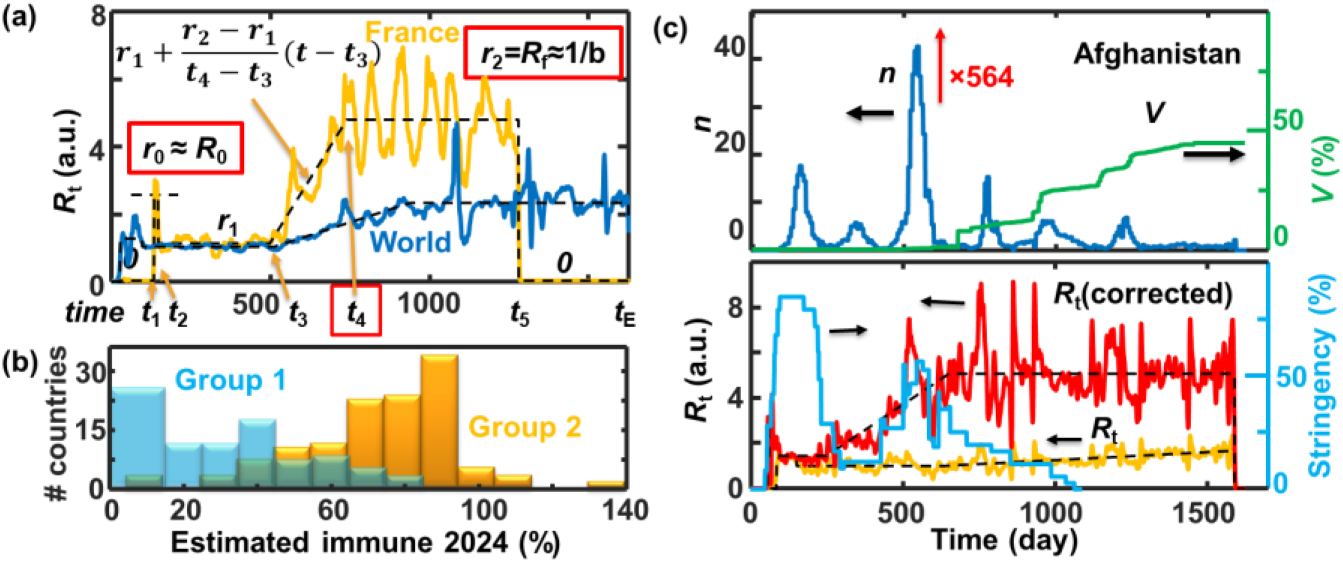
(a) Example fitting of *R*_t_ curve using the laydown ‘5’ shape for France and world average. (b)Histogram of Group 1 countries that do not follow (a) and Group 2 countries that follow the laydown ‘5’ pattern in (a) with today’s immune ratios. (c) Example data treatment of Group 1 countries with low COVID-19 testing rates, briefly, multiplying a testing rate correction factor estimated using world average timing and recalculating the *R*_t_ values.

In the next step, we manually check the *R*_t_ curves of the 254 records in the raw data to group them into three groups: 87 countries that do not follow (Group 1), 121 countries follow the laydown ‘5’ pattern (Group 2), and the rest, being the averages of groups of countries, or countries missing a significant amount of data.

Simplifying the total immune population as cumulated *n* times *γ* plus cumulative fully vaccinated population,

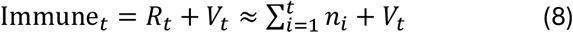

The 87 Group 1 countries have a relatively low average immune population (0.30±0.20) compared to that of the Group 2 countries (0.73±0.21) at the end date of the raw data accessed in July 2024 (**Figure 6b**). These values are correlated with GDP per capita, which makes sense because both testing and vaccination cost money. The average GDP per capita of Group 1 countries is $6000, and Group 2 countries is $20,000. Many Group 1 countries have relatively low testing rates, thus a smaller *R*_t_ at the later phase in the SIRVB model compared to Group 2 countries. Some countries also have low vaccination rates. To simplify our analysis, we assume that at the global herd immunity day, these countries also reach herd immunity with an assumed ∼67% immune population. We assume a constant correction factor to be,

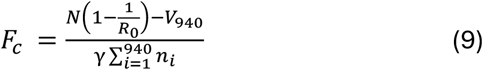

And the corrected new case per day is

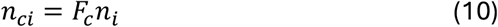

The average correction factor for the 87 Group 1 countries is 775 (1-9000), suggesting a very low testing ratio for many countries.

After the daily case correction, the *R*_t_ values are recalculated, and a laydown 5 shape is observed for most of the Group 1 countries. For example, the first country on the list, Afghanistan, has a low detected cases per day per million people, peaking ∼40 (**Figure 6c**). It also has relatively low vaccination at ∼20% around day 940. Thus, its *R*_t_ is almost flat over time. This data suggests it is still at an early phase till the last day of the raw data in the SIRVB model, which is unlikely true. We apply a correction factor of 564 to the data and a new *R*_t_ curve showing a laydown 5 shape. Its trend anti-correlates with the stringency index very well. The herd-immunity date for Afghanistan is re-defined to the 620^th^ day on the revised *R*_t_ curve, right after the last major social restriction peak and the most intensive outbreak peak (**Figure 6c**).

After we have re-defined the herd immunity date for the Group 1 countries in the model, we correlate these dates with other social parameters to check for a pattern among them (**Figure 7**). We leave the Group 2 countries with low testing ratio unchanged, resulting in some having a later herd-immunity date than the rest of the countries. The average day to reach herd immunity (SIRVB model) for Group 1, mostly developing countries, is 655±184 days, and for Group 2, including most developed countries is 873±134 days. After analyzing the correlations of the herd-immunity days with other parameters (**Figure 7**), we suspect this difference is due to the two different strategies of achieving herd immunity, mainly through infection *vs* mainly through vaccination. Also, many developing countries have lower median ages ^42^. Because COVID-19 is less toxic to young people ^43^, choosing infection to gain herd-immunity for young people is a natural choice.

**Figure 7.**
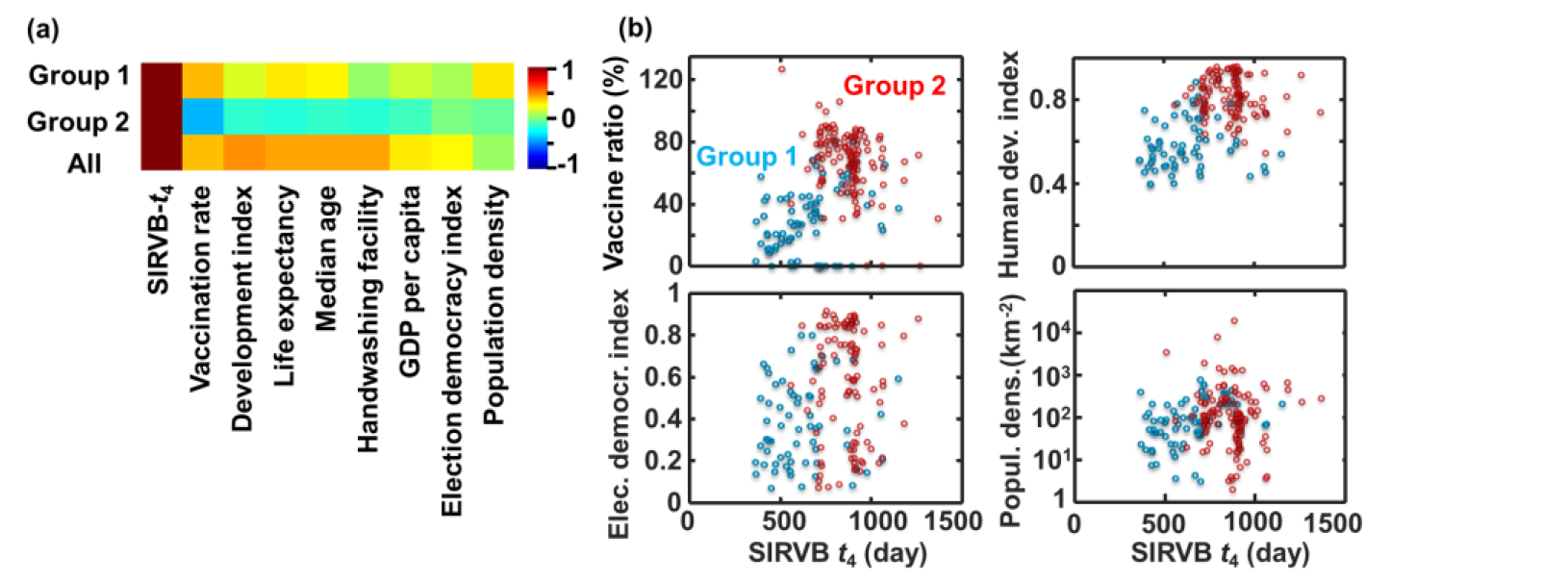
(a) Correlation of the turning time *t*_4_ in the SIRVB model with parameters of countries from Our World in Data. (b) Example plots of *t*_4_ with four parameters. Fall data available in the **SI**.

A few correlations are further expanded in **Figure 7b**. Our assigned herd-immunity time (*t*_4_ in **Figure 6**) is positively correlated with vaccination for Group 1 and all countries. This correlation makes sense because *I* and *V* determine *t*_4_ in the SIRVB model. Interestingly, a negative correlation is observed within Group 2 countries. We suspect it to be the cost of time to wait for the vaccination to be available. Group 1 countries have significantly shorter time than Group 2 because they achieve herd-immunity mainly via infection with an average median age of 24±8; thus, there is less need to wait for the vaccines as Group 2 countries with an average median age of 36±7. Average population density should not matter because it is the local or median population density that determines the *R*_0_ and *R*_t_ values. Thus, a significant correlation between this parameter and the herd-immunity day is not observed for all countries. The median age, life expediency, handwashing facility, and election democracy index all contribute to the calculation of the human development index. Thus, their distribution *vs t*_4_ shows a positive correlation with a binary pattern between the two groups.

## Conclusions

We believe the SIRVB model is the simplest mathematical model to explain the COVID-19 kinetic data of all countries in Our World in Data (raw data) from the World Health Organization (WHO). The calculated *R*_t_ values from the model are strongly anti-correlated with the stringency index estimated in the raw data. Thus, it is possible to calculate the *R*_t_ values of the SIRVB model just using stringency-index and *R*_0_ in a future outbreak of an epidemic or pandemic, providing valuable prediction power to researchers and policymakers, especially in regions where testing is rare.

In summary, there is an epidemiological significance of SIR, SIRV, and SIRVB models, and choosing a proper model and finding out the proper parameters could be essential in predicting trends and suggesting policy changes for a future epidemic or pandemic. A modification of the SIRVB model allows the adjustment of low-statistics data to obtain otherwise hidden *R*_t_ values, taking advantage of parameters fitted from the data of high-statistics regions. Thus, the central disease monitoring department, such as WHO or the disease control department of a country can have an estimation of the stages of the spreading in low statistics regions.

A few weak points of the simplified SIRVB model may need further improvement to a more sophisticated model. The post-pandemic data analysis we have carried out is a mathematical practice based on naïve assumptions rather than a restricted epidemiological study. We believe the parameters of the SIRVB model can be obtained experimentally during a pandemic and used to predict its trend. Due to our limited knowledge of epidemiology, we will leave this task to others who are more suitable. The model does not have an internal overlap of populations; neither does the model have imported cases. Time-dependent variables such as *γ* and *b* are assumed constants. Estimation of testing and vaccination rates have significant uncertainties. Also, the model treats the whole entity, either a city, a state, a country, or the whole world, uniformly. These assumptions introduce errors in *R*_t_ calculation, which is also part of why the after-pandemic *R*_t_ approaches 1/*b* rather than (should be) *R*_0_. The number of susceptible populations will be estimated to be 0 at a later phase, and all cases are breakthrough which is not true. Thus, any tested positive case, even 1 in the whole population, will give a calculated *R*_t_ = 1/*b* losing its significance.

## Data Availability

All data produced in the present work are contained in the manuscript.

## ASSOCIATED CONTENT

Supporting Information

Data is available in Excel sheets for the figures in this report.

## AUTHOR INFORMATION

### Data accessibility

All raw data is downloaded from Our World in Data that is publicly accessible. Data in figures are also included in the supporting information. MATLAB (2024a) source code will be uploaded to GitHub that is publicly accessible. Thus, all results in this paper should be reproducible.

## ACKNOWLEDGMENTS

We thank the National Human Genome Research Institute of the National Institutes of Health (award number 2R15HG009972) for supporting our teaching practices. The content is solely the responsibility of the authors and does not necessarily represent the official views of the National Institutes of Health.

